# Environmental injustice - Neighborhood characteristics as confounders and effect modifiers for the association between air pollution exposure and cognitive function

**DOI:** 10.1101/2021.10.19.21265212

**Authors:** Zhenjiang Li, Grace M. Christensen, James J. Lah, Michele Marcus, Armistead G. Russell, Stefanie Ebelt, Lance A. Waller, Anke Huels

**Affiliations:** Gangarosa Department of Environmental Health, Rollins School of Public Health, Emory University, Atlanta, Georgia, USA; Department of Epidemiology, Rollins School of Public Health, Emory University, Atlanta, Georgia, USA; Department of Neurology, School of Medicine, Emory University, Atlanta, Georgia, USA; School of Civil and Environmental Engineering, Georgia Institute of Technology, Atlanta, Georgia, USA; Department of Biostatistics and Bioinformatics, Rollins School of Public Health, Emory University, Atlanta, Georgia, USA

## Abstract

**Background:** Air pollution has been associated with cognitive decline among the elderly. Previous studies have not evaluated the simultaneous effect of neighborhood-level socioeconomic status (N-SES), which can be an essential source of bias.

**Objectives:** We explored N-SES as a confounder and effect modifier in a cross-sectional study of air pollution and cognitive function among the elderly.

**Methods:** We included 12,058 participants age 50+ years from the Emory Healthy Aging Study in Metro Atlanta using the Cognitive Function Instrument (CFI) score as our outcome, with higher scores representing worse cognition. We estimated 9-year average ambient carbon monoxide (CO), nitrogen oxides (NO_x_), and fine particulate matter (PM_2.5_) concentrations at residential addresses using a fusion of dispersion and chemical transport models. We collected census-tract level N-SES indicators and created two composite measures using principal component analysis and k-means clustering. Associations between pollutants and CFI and effect modification by N-SES were estimated via linear regression models adjusted for age, education, race and N-SES.

**Results:** N-SES confounded the association between air pollution and CFI, independent of individual characteristics. We found significant interactions between all air pollutants and N-SES for CFI (*p*-values<0.001) suggesting that effects of air pollution differ depending on N-SES. Participants living in areas with low N-SES were most vulnerable to air pollution. In the lowest N-SES urban areas, interquartile range (IQR) increases in CO, NO_x_, and PM_2.5_ were associated with 5.4% (95%-confidence interval, -0.2,11.4), 4.9% (-0.4,10.4), and 9.8% (2.2,18.0) increases in CFI, respectively. In lowest N-SES suburban areas, IQR increases in CO, NO_x_, and PM_2.5_ were associated with higher increases in CFI, namely 13.4% (1.3,26.9), 13.4% (0.3,28.2), and 17.6% (2.8,34.5), respectively.

**Discussion:** N-SES is an important confounder and effect modifier in our study. This finding could have implications for studying health effects of air pollution in the context of environmental injustice.

## INTRODUCTION

During the past decade, multiple investigations explored the effects of long-term exposure to air pollution on cognitive function in the older population after animal models had shown effects of air pollution on the central nervous system via inflammation and oxidative stress (Block & Calderon-Garciduenas, 2009). Delgado-Saborit et al. reviewed 69 relevant epidemiological studies from 2006 to 2019 and found consistent associations of long-term exposure to air pollution with reduced global cognition, cognitive decline, and dementia (Delgado-Saborit et al., 2021).

Individuals living in disadvantaged neighborhoods are often exposed to the highest concentrations of air pollution – a problem known as environmental injustice (Maantay, 2002). Exposure to air pollution is considered to interact with vulnerability, producing a “triple jeopardy” of low socioeconomic position, polluted environment, and impaired health (J. Ailshire, Karraker, & Clarke, 2017; Hicken et al., 2016; Ou et al., 2008). Neighborhoods have emerged as a common analysis unit to identify disadvantaged populations in studies of environmental injustice. A variety of epidemiological studies conducted among different racial/ethnic groups throughout the world have demonstrated associations between living in low N-SES neighborhoods and cognitive decline in the elderly, associations independent of individual-level characteristics (Basta, Matthews, Chatfield, Brayne, & Mrc, 2008; Espino, Lichtenstein, Palmer, & Hazuda, 2001; Guo, Chan, Chang, Liu, & Yip, 2019; Lang et al., 2008; Sheffield & Peek, 2009; Shih et al., 2011; Wee et al., 2012; Wight et al., 2006). Therefore, there is a strong likelihood that neighborhood characteristics may confound and/or modify the association between air pollution and cognitive functioning. However, only a handful of previous studies on air pollution and cognitive function controlled for neighborhood educational attainment, racial composition, and socioeconomic context (J. A. Ailshire & Crimmins, 2014; Bowe, Xie, Yan, & Al-Aly, 2019; Cullen et al., 2018), and only one study included neighborhood psychological stressors as potential effect modifiers (J. Ailshire et al., 2017).

In the current study, we conducted a cross-sectional analysis to assess the confounding and modifying effect of neighborhood-level socioeconomic status (N-SES) in exploring the associations between long-term exposure to ambient air pollution [carbon monoxide (CO), nitrogen oxides (NO_x_), and fine particulate matter (PM_2.5_)] and cognitive function among residents over 50 years of age living in Metro Atlanta (i.e., Atlanta metropolitan area, the ninth-largest metropolitan statistical area in USA), GA, USA. For assessing N-SES, we used a combination of N-SES composite measures at different geographic levels. Our specific objectives were to determine whether (1) long-term exposure to ambient air pollution adversely affects cognitive function in older population after adjusting for individual and neighborhood-level confounders; (2) individuals living in disadvantaged neighborhoods are more susceptible to the effect of air pollution on cognitive function.

## METHODS

### Participants and Study Design

The present study used a cross-sectional assessment of baseline data from the Emory Healthy Aging Study (EHAS), an on-going population-based cohort that was launched in 2015 (Goetz et al., 2019; Wingo et al., 2020). Briefly, the EHAS aims to better understand factors contributing to healthy aging and identify markers that can predict common age-related diseases. Participants are enrolled from the population who receive health services at Emory Healthcare (headquartered in Atlanta, GA, USA), as well as their spouses, family members and associated non-relatives. Information is collected through an online Health History Questionnaire (HHQ), which contains items on demographics, general heath, mental and cognitive health, and participant contact information. In addition, all EHAS participants are invited to complete remote cognitive assessments through the EHAS mobile application. All participants complete an online consent process prior to enrollment, and the study was approved by the Emory University Institutional Review Board.

### Assessment of Cognitive Function

The Cognitive Function Instrument (CFI) was used to assess cognitive status of participants at baseline, with higher scores reflecting more perceived memory decline and cognitive decline (Amariglio et al., 2015; C. Li et al., 2017). The CFI has 14 items that efficiently probe the full realm of subjective cognitive concerns interfering with the daily activities of older adults. Previous studies demonstrated that the CFI was a sensitive and reliable instrument in tracking early decline in cognitive function in older adults. Total scores on the instrument range from 0 to 14, and response to each item was based on a 1-point scale (Yes = 1, No = 0, and Maybe = 0.5).

### Assessment of Exposure

A data fusion approach that combines Research Line (R-LINE) source model and Community Multiscale Air Quality (CMAQ) model simulations, along with observations, was used to estimate long-term exposure to ambient CO, NO_x_, and PM_2.5_ at each participant’s geocoded residential address for a fixed 9-year period (2002-2010). The development of the fusion model has been described elsewhere (Senthilkumar et al., 2019; Yu et al., 2018). Briefly, R-LINE is a dispersion model for near surface emissions from line sources, such as roadways, and was applied to produce a 250 m grid resolution field, and CMAQ is a chemical transport model applied at a 12 km grid resolution to provide regional influences. The fusion model, which takes advantage of both air quality models, generated daily concentration estimates from 2002 to 2010 of 1-hr CO, NO_x_, and 24-hr average PM_2.5_ at a 250 m grid resolution across Metro Atlanta. We spatially matched geocoded residential addresses to the closest centroid of grids, and calculated the individual long-term exposure as the average concentration from 2002 to 2010 of the matched grids. We divided the averaged concentration by the corresponding interquartile ranges (IQR) of air pollutants (CO, 328.4 ppb; NO_x_, 28.0 ppb; PM_2.5_, 1.3 µg/m^3^) to enable comparison among pollutant-specific effect estimates from epidemiologic models.

### Assessment of Neighborhood-level Socioeconomic Status

We obtained estimates of census-tract-level SES from the American Community Survey (ACS) 5-year summary via R package *tidycensus* 0.10.2 (Walker, Eberwein, & Herman, 2018). Census tracts are fairly homogenous units and have acceptable data completeness, and area-based socioeconomic measures at this geographic level were appropriate for U.S. public health and research (Krieger et al., 2002). To achieve a good overall representation of neighborhood-level SES, we generated composite measures based on 16 indicators corresponding to six socioeconomic domains (poverty/income, racial composition, education, employment, occupation, and housing properties), which have been widely used in previous studies of SES and health (Messer et al., 2006). Poverty variables included percent households in poverty, percent female headed households with dependent children, percent households earning under $35,000 per year, percent households on public assistance, and percent households with no car. Racial composition was estimated using percent residents who were non-Hispanic blacks. Education included percent males and females with less than a high school education. Employment variables included unemployment rate and percent males no longer in work force. Occupation variables included percent males and females not in management, business, science, and arts occupations. Housing variables included percent rented, percent vacant, percent crowded households, and median household value. We also defined an indicator for residential stability by calculating the estimated percent of households moved in the current residence before 2010 based on the ACS for sensitivity analysis. Geocoded residential addresses were mapped to the census tract reference map published by U.S. Census Bureau. As the enrollment year of participants ranged from 2015 to 2020 in our EHAS dataset, and the most recent data release from the ACS is 2018, participants were matched to ACS datasets 2 years prior to their enrollment. We employed two dimension reduction approaches to leverage the information captured by the 16 N-SES indicators: principal component analysis (PCA) and k-means clustering analysis (KMCA). We used KMCA to generate clusters of census tracts with similar characteristics, and used the Elbow method to determine the optimal number of clusters (Figure S1B) (Kodinariya & Makwana, 2013). We used PCA to generate census-tract-level PCs (Abdi & Williams, 2010), which captured an additional layer of differences in N-SES within these clusters. We included principal components (PCs) that explained at least 80% of the total variance in the models (Figure S1A). Briefly, we found that KMCA generated a larger geographic unit of neighborhoods with relatively similar characteristics and the PCs captured an additional layer of differences in N-SES within these clusters (Figure S2).

### Assessment of Individual-level Characteristics

We derived individual-level characteristics of participants from the EHAS online questionnaire, and included age, sex, race, Hispanic ethnicity, household income, education attainment, and disease history of mild cognitive impairment, Alzheimer’s disease, or other dementias. We classified racial identification into White, Black, and others that included American Indian or Alaska Native, Asian, and Native Hawaiian or other Pacific Islander. We note that the US Census includes a separate question on Hispanic ethnicity, participants who self-reported as of Hispanic origin also select race from the categories above. We classified continuous annual income into 3 categories: less than $50,000, $50,000-100,000, and more than $100,000. We defined individual education attainment as the highest degree participants reported receiving and categorized values into the following categories: high school or less, some college credit, associate degree, bachelor degree, master degree, and professional or doctorate degree.

### Statistical Analysis

We summarized characteristics of the total study population and stratified by quartiles of CFI to describe the potential patterns of covariates in dependence of CFI. We present the distribution of long-term exposure concentrations of ambient air pollutants and the individual N-SES characteristics for each N-SES cluster as defined by KMCA.

We tested the associations between air pollutants and CFI using multiple linear regression models taking the natural log of CFI as the dependent variable due to the skewed distribution of CFI values. Due to the potentially complex confounding structure (Figure S4), we used a stepwise procedure to assess the association of air pollution and CFI, and then the impact of confounding by N-SES on the association between air pollution and CFI. We added two composite measures of N-SES into the models separately or together to assess the confounding of N-SES at different geographic scales. The following single-pollutant models were fitted using adjusted linear regression analyses:

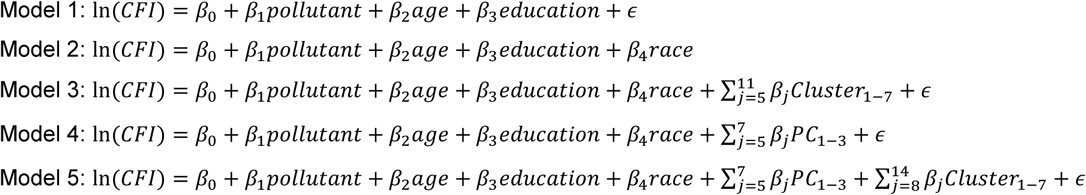

where In(*CFI*) refers to the natural log-transformed CFI [a constant (0.5) was added to handle zero values prior to applying the log transformation] due to the skewed distribution of raw CFI (Figure S3); *pollutant* and *age* were included as continuous variables; *education* was a categorical variable with six levels (high school or less, some college credit, but no degree, associate degree, bachelor’s degree, master’s degree, professional or doctorate degree); *cluster*_1-7_ refers to the KMCA clusters derived from N-SES indicators; *PC*_1-3_ refers to the first three principal components derived from N-SES, which explained at least 80% of the total variance.

The model series (Model 1-5) evaluated variables anticipated to confound the association of air pollution with cognitive function based on a conceptualized directed acyclic graph (DAG, Figure S4). The minimal sufficient set of confounders identified by the DAG contained age, individual-level SES, and N-SES. As many of our older study participants were likely retired, we used educational attainment as a measure of individual-level SES instead of income. Individual race was added in Models 2-5 to control for potential individual-SES confounding that may not be captured by education alone (Figure S4). We further included N-SES PCs and clusters in Models 3-5, to evaluate whether N-SES was a confounder for the association between air pollutants and CFI in addition to individual-level SES. N-SES was represented by two composite measures at different geographic levels in our study, and we assessed the existence of residual confounding by N-SES by adding the two composite measures into the models individually (Models 3 and 4) and simultaneously (Model 5).

To analyze potential effect modification by N-SES on the association between a single pollutant and CFI, we added a product term of *cluster*_1-7_ and a *pollutant* to Model 5, as showed below:

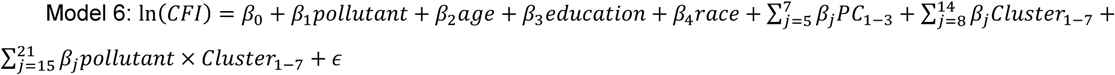

We calculated the cluster-specific coefficients for air pollution by summing up the coefficients of *pollutant* and the product term except for the referent cluster in which case we used the coefficient of *pollutant* itself. To investigate the robustness of the results of the interaction analysis to the length of time participants may have been living at their current residence (unknown in our current cohort data), we performed a sensitivity analysis in which we included a neighborhood-level indicator of residential stability as an additional covariate in the model.

We performed the spatial join of geocoded residential address, 250 m grid cells of air pollution data, and census tract SES data using ArcGIS Pro version 2.5.2 (Esri, Redlands, CA, USA), and statistical analyses were performed using R version 4.0.3 (R Core Team). Statistical significance was set at *p*-value < 0.05.

## RESULTS

### Study Characteristics

Until the first quarter of 2020, the EHAS cohort had enrolled 20,523 participants from all over the USA, with 14,404 participants living in Metro Atlanta at enrollment. The current study included all participants over 50 years old at enrollment living in Metro Atlanta, resulting a final sample size of 12,058 (Table 1). The average age of the study participants was 64 years. About 70% of the participants were female, and this proportion was similar in the original cohort without any exclusion (Goetz et al., 2019). Only about 10% of participants were African Americans (AA), and most of participants had relatively high socioeconomic status based on their household income and educational attainment. For example, almost 50% of the study population had an annual household income more than $100,000, given the median household income in Atlanta was $59,948. Over 70% of the study population had a bachelor’s degree or higher, while about 50% for the general population. The majority of participants were free of mild cognitive impairment (MCI), Alzheimer’s disease (AD), or other dementias. The mean age was marginally higher in the third (Q_2_ – Q_3_ CFI) and fourth quartiles of the CFI distribution (Q_3_ – Max CFI). The first quartile had the highest proportion of White, while the fourth quartile had the highest proportion of racial minorities. In addition, participants with a higher CFI were more likely to have a lower income and education (Figure S5).

**Table 1.** Study characteristics stratified by quartiles (Q_1_ to Q_3_) of cognitive function (cognitive function instrument, CFI).

| Characteristics | Total<br>(N = 12,058) | Min - Q<sub>1</sub> CFI <sup>a</sup><br>(N = 3,280) | Q<sub>1</sub> - Q<sub>2</sub> CFI<br>(N = 2,879) | Q<sub>2</sub> - Q<sub>3</sub> CFI<br>(N = 3,089) | Q<sub>3</sub> - Max CFI<br>(N = 2,810) |
| --- | --- | --- | --- | --- | --- |
| Age |  |  |  |  |  |
| Mean (SD) | 65.0 (8.40) | 64.4 (8.15) | 64.8 (8.15) | 65.6 (8.33) | 65.0 (8.96) |
| Gender |  |  |  |  |  |
| Female | 8,490 (70.5%) | 2201 (67.2%) | 2055 (71.5%) | 2219 (71.9%) | 2015 (71.8%) |
| Male | 3,553 (29.5%) | 1072 (32.8%) | 819 (28.5%) | 869 (28.1%) | 793 (28.2%) |
| Missing | 15 | 7 | 5 | 1 | 2 |
| Race |  |  |  |  |  |
| White/Caucasian | 9,778 (81.9%) | 2741 (84.3%) | 2372 (83.1%) | 2534 (82.9%) | 2131 (76.7%) |
| Black/African American | 1,587 (13.3%) | 382 (11.8%) | 356 (12.5%) | 394 (12.9%) | 455 (16.4%) |
| Others <sup>b</sup> | 573 (4.8%) | 127 (3.9%) | 126 (4.4%) | 129 (4.2%) | 191 (6.9%) |
| Missing | 120 | 30 | 25 | 32 | 33 |
| Hispanic |  |  |  |  |  |
| No | 11624 (96.6%) | 3175 (96.9%) | 2785 (96.9%) | 2985 (96.8%) | 2679 (95.5%) |
| Yes | 414 (3.4%) | 100 (3.1%) | 89 (3.1%) | 98 (3.2%) | 127 (4.5%) |
| Missing | 20 | 5 | 5 | 6 | 4 |
| Household income (\$) | | | | | |
| USD < 50,000 | 2,075 (18.1%) | 407 (13.1%) | 414 (15.1%) | 523 (17.7%) | 731 (27.1%) |
| USD 50,000-100,000 | 3,859 (33.6%) | 936 (30.2%) | 885 (32.3%) | 1056 (35.8%) | 982 (36.4%) |
| USD >= 100,000 | 5,545 (48.3%) | 1753 (56.6%) | 1437 (52.5%) | 1368 (46.4%) | 987 (36.6%) |
| Missing | 579 | 184 | 143 | 142 | 110 |
| Highest education |  |  |  |  |  |
| High school or less | 482 (4.0%) | 98 (3.0%) | 103 (3.6%) | 114 (3.7%) | 167 (6.0%) |
| Some college credit, but no degree | 1,885 (15.7%) | 450 (13.7%) | 419 (14.6%) | 471 (15.3%) | 545 (19.4%) |
| Associate degree | 877 (7.3%) | 198 (6.0%) | 188 (6.5%) | 204 (6.6%) | 287 (10.2%) |
| Bachelor degree | 4,087 (33.9%) | 1081 (33.0%) | 1003 (34.9%) | 1061 (34.4%) | 942 (33.6%) |
| Master degree | 3,097 (25.7%) | 895 (27.3%) | 778 (27.1%) | 822 (26.7%) | 602 (21.5%) |
| Professional or doctorate degree | 1,615 (13.4%) | 556 (17.0%) | 385 (13.4%) | 411 (13.3%) | 263 (9.4%) |
| Missing | 15 | 2 | 3 | 6 | 4 |
| CFI |  |  |  |  |  |
| Median (IQR) | 1.5 (2.5) | 0 (0.5) | 1 (0.5) | 2.5 (1) | 5 (2.5) |
| Mild cognitive impairment |  |  |  |  |  |
| No | 11,144 (97.1%) | 3137 (99.3%) | 2736 (99.1%) | 2879 (98.0%) | 2392 (91.4%) |
| Yes | 333 (2.9%) | 23 (0.7%) | 26 (0.9%) | 60 (2.0%) | 224 (8.6%) |
| Missing | 581 | 120 | 117 | 150 | 194 |
| Alzheimer's disease |  |  |  |  |  |
| No | 11,431 (99.3%) | 3145 (99.5%) | 2755 (99.5%) | 2941 (99.5%) | 2590 (98.7%) |
| Yes | 79 (0.7%) | 17 (0.5%) | 14 (0.5%) | 15 (0.5%) | 33 (1.3%) |
| Missing | 548 | 118 | 110 | 133 | 187 |
| Other dementia |  |  |  |  |  |
| No | 11,366 (99.3%) | 3139 (99.6%) | 2742 (99.5%) | 2917 (99.6%) | 2568 (98.5%) |
| Yes | 77 (0.7%) | 14 (0.4%) | 13 (0.5%) | 12 (0.4%) | 38 (1.5%) |
| Missing | 615 | 127 | 124 | 160 | 204 |

<sup>a</sup> The study population was stratified by the quartiles of CFI (Q<sub>1</sub>, 0.5; Q<sub>2</sub>, 1.5; Q<sub>3</sub>, 3.0). Higher values of CFI correspond to worse cognitive functioning.
<sup>b</sup> Others included Asian, American Indian, Alaska Native, Native Hawaiian and other pacific islander, and multi-race.

Figure 1 summarizes the information on N-SES. Based on the 16 N-SES indicators, KMCA identified seven clusters (Figure 1A). We determined the optimal number of clusters via total intra-cluster variation calculated by the Elbow method (Figure S1B). Clusters 3, 4, 6, and 7 were the most distinguishable based on extreme values in at least one socioeconomic characteristic or a specific spatial pattern (Figure 1A & 1C). Cluster 6 comprised primarily central areas of Atlanta with the highest proportion of non-Hispanic Black population. Cluster 7 covered a smaller area mainly around Buford Highway northeast of the city which is home to a concentration of Hispanic and immigrant communities. The Clusters 6 & 7 also represent some of the lowest N-SES neighborhoods in Metro Atlanta as indicated by unemployment rate, percent individuals in poverty, percent households on public assistance, percent individuals with education less than the high school and percent crowded households. About half of the residents in both clusters had an annual household income less than $35,000, which is less than 1.5 times of the 2021 federal poverty line for families of four (USDHHS, 2020). In contrast, Cluster 4 represented the highest N-SES. Cluster 3 consisted of census tracts along major highways in northern Atlanta. As for ambient air pollution, participants living in clusters with a lower level of N-SES (C6 and C7) were generally exposed to higher concentrations of air pollution at their residences; Clusters 6 and 7 had the second and third highest median air pollution concentrations. For those living in Cluster 3, residential air pollution exposures were also high due to the proximity to highways.

**Figure 1.**
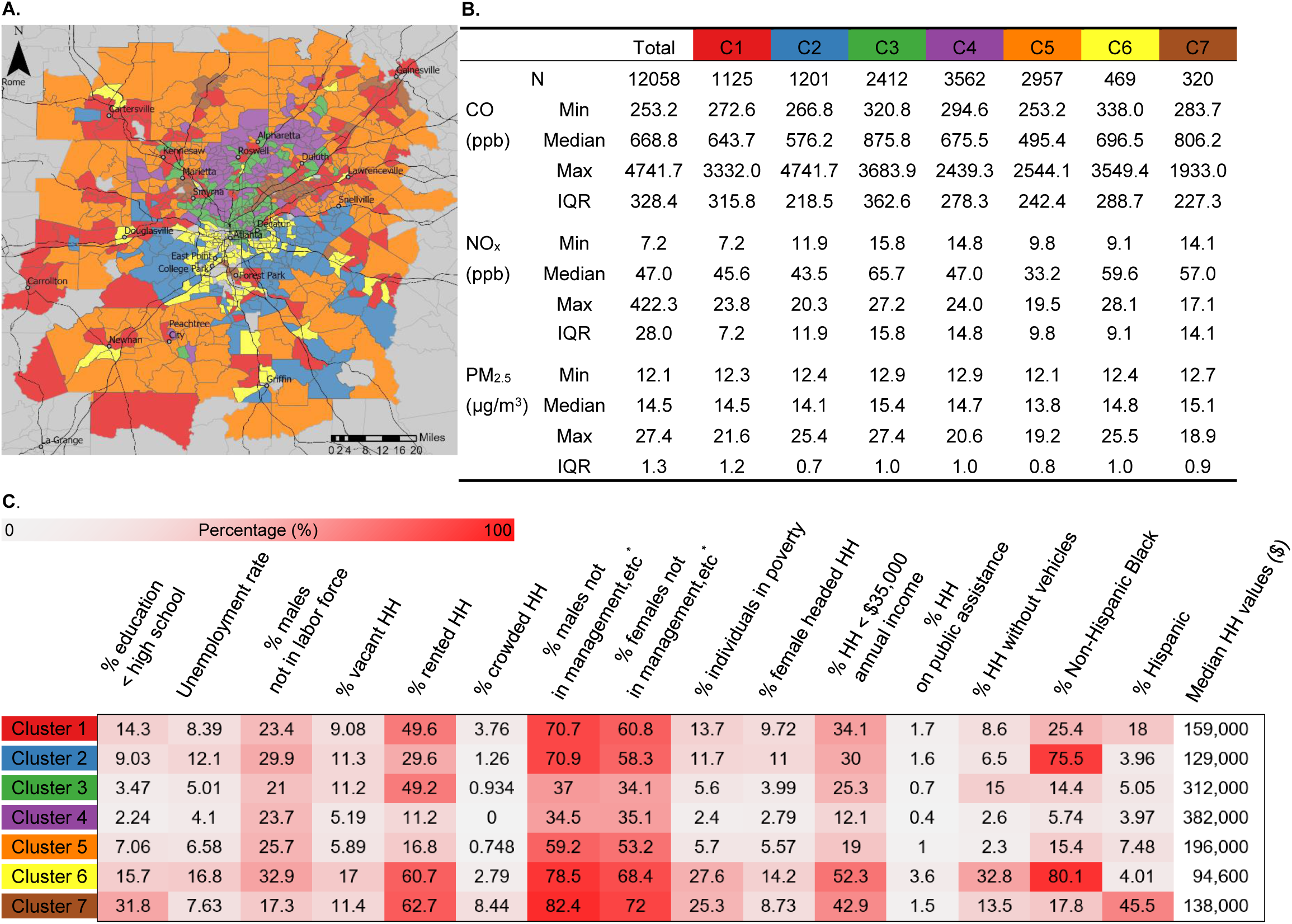
Statistics of Air pollution concentrations and neighborhood socioeconomic indicators for the clusters of census tracts. **A**. Seven clusters generated by k-means cluster analysis in Metro Atlanta. Highways were denoted by solid black lines, and the gray lines depicted the boundaries of census tracts. **B**. The distribution of individual exposures to ambient air pollution for each cluster. **C**. Neighborhood socioeconomic status (N-SES) of each cluster described by the median of 16 N-SES indicators at census-tract level. Abbreviations: N, number of participants; C1-7, Cluster 1-7; CO, carbon monoxide; NO_x_, nitrogen oxides; PM_2.5_, fine particulate matters; IQR, interquartile range; HH, households; management, etc. included management, business, science, and arts occupations.

### Air pollution, N-SES and cognitive functioning

When assessing associations of air pollution and CFI, we observed that higher exposure to ambient PM_2.5_ was associated with better cognitive function when only adjusting for age, individual-level education, and race [e.g., -2.2% (95% CI, -3.9, -0.4%) CFI per 1.3 µg/m^3^ increase in PM_2.5_] (Figure 2A, Models 1 and 2). Adding the N-SES clusters and PCs as confounding factors attenuated the association towards the null [Model 3: -1.5% (-3.5, 0.58%); Model 4: 0.3% (-2.1, 2.6%); Model 5: 0.4% (-1.9, 2.8%)]. As such, ignoring N-SES as a confounding factor resulted in biased effect estimates and an erroneous significantly inverse association between PM_2.5_ and CFI.

**Figure 2.**
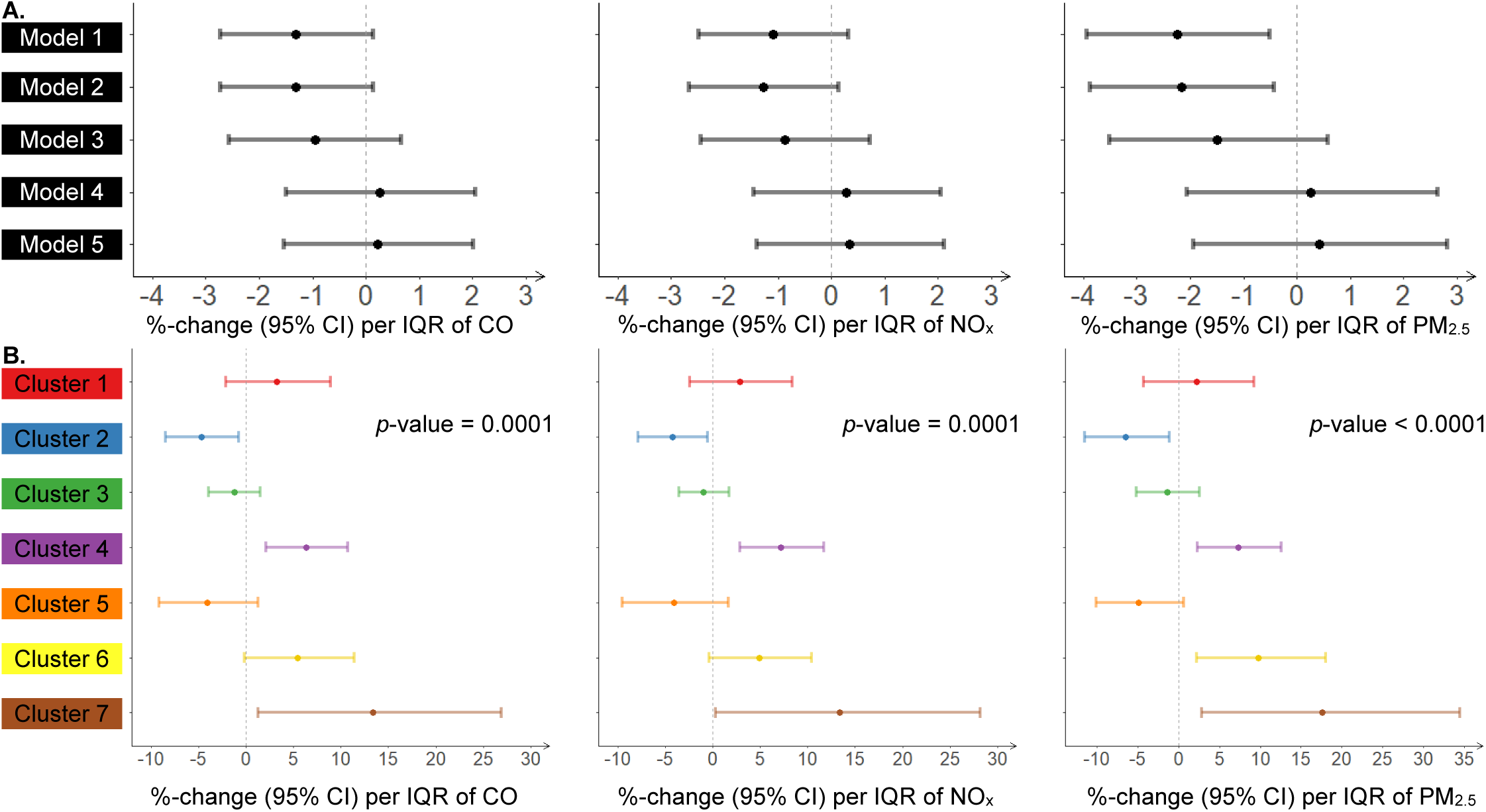
Associations between 9-year averaged exposure to ambient air pollution and Cognitive Function Instrument (CFI). Percent change (%) of CFI associated with an interquartile range (IQR) increase (CO, IQR=328.4ppb; NO_x_, IQR=28.0ppb; PM_2.5_, IQR=1.3µg/m^3^) in the exposure estimate and corresponding 95% confidence intervals are presented. **A**. Confounding by N-SES. Model 1: adjusted for age and individual educational attainment; Model 2: adjusted for age, individual educational attainment, and race; Model 3: adjusted for covariates in Model 2 and N-SES clusters; Model 4: adjusted for covariates in Model 2 and N-SES PCs; Model 5 (main model): adjusted for covariates in Model, N-SES clusters and PCs simultaneously. **B**. Effect modification by N-SES. Associations between 9-year averaged exposure to ambient air pollution and Cognitive Function Instrument stratified by the N-SES cluster the participants were assigned to (compare Figure 1A). Associations were adjusted for age, individual educational attainment, race, and N-SES PCs at census-tract level. The significance of interaction was indicated by *p*-values in each plot. Note: CO, carbon monoxide; NO_x_, nitrogen oxides; PM_2.5_, fine particulate matter.

In addition, we found effect modification by N-SES clusters for the association between air pollution and cognitive functioning, with statistically significant interaction terms for all three air pollutants (CO, *p* = 0.0001; NO_x_, *p* = 0.0001; PM_2.5_, *p* < 0.0001; Figure 2B and Table S2). The strongest association between air pollution and cognitive functioning was found in participants assigned to N-SES Cluster 7, the cluster characterized by a high proportion of Hispanic and immigrant communities and high air pollution concentrations. Among participants living in Cluster 7, IQR increases in CO, NO_x_, and PM_2.5_ were associated with 13.4% (95% CI, 1.3, 26.9%), 13.4% (0.3, 28.2%), and 17.6% (2.8, 34.5%) increases of CFI, respectively. Among participants living in Cluster 6, comprising primarily central areas of Atlanta with the highest proportion of non-Hispanic Black population, IQR increases in CO, NO_x_, and PM_2.5_ were associated with 5.4% (95% CI, -0.2, 11.4%), 4.9% (-0.4, 10.4%), and 9.8% (2.2, 18.0%) increases of CFI, respectively. Among participants living in Cluster 4, the cluster with the highest N-SES, IQR increases in CO, NO_x_, and PM_2.5_ were associated with 6.3% (95% CI, 2.1, 10.8%), 7.2% (95% CI, 2.9, 11.7%), and 7.3% (95% CI, 2.2, 12.6%) increases of CFI, respectively. We did not observe significant associations of air pollution and CFI for Clusters 3 or 5. Cluster 2 was the only cluster in which we found an inverse association between air pollution and cognitive functioning. Neighborhoods assigned to Cluster 2 were characterized by the second highest proportion of non-Hispanic Black population and intermediate N-SES.

As population mobility may impact the exposure assessment, we used the neighborhood-level indicator of residential stability as a surrogate for the length of time participants may have been living at their current residence (unknown in our current cohort data) and investigated the robustness of the results of the interaction analysis to it. The results of sensitivity analyses are summarized in Figure S6. All associations and effect modifications were robust to additional adjustment for residential stability.

## DISCUSSION

In the present study of 14,404 individuals 50 years and older from Metro Atlanta, we observed a significant effect modification by N-SES for the association between air pollution and cognitive function, and significant associations between air pollution exposure and cognitive decline among participants living in the most socioeconomically disadvantaged neighborhoods. In addition, our study demonstrates the importance of including N-SES as confounding variable when analyzing associations between air pollution and cognitive function. Not adjusting for N-SES resulted in a significant negative association between the long-term exposure to air pollution and CFI scores, which indicated a likely “erroneous” protective effect of air pollution, despite adjusting for individual age, education attainment, and race. These findings show the importance of including neighborhood characteristics as confounders and effect modifiers when analyzing associations between air pollution and cognitive function.

A wealth of epidemiological studies has shown an association of long-term exposure to ambient PM_2.5_ with the risk of dementia (Fu, Guo, Cheung, & Yung, 2019). Although the use of various cognitive assessment tools and exposure metrics have made it hard to compare the effect estimates across different studies, a higher exposure to PM_2.5_ has been consistently associated with worse global cognition independent of individual demographics in most previous studies (J. Ailshire et al., 2017; J. A. Ailshire & Crimmins, 2014; Lin et al., 2017; Shin, Han, & Choi, 2019; Tallon, Manjourides, Pun, Salhi, & Suh, 2017). In addition, including area-level socioeconomic features, such as the Townsend score in the UK, often attenuated observed associations towards the null (Cullen et al., 2018; Tallon et al., 2017). In the current study, we observed a significant protective effect of PM_2.5_ on cognitive function when adjusting for individual demographics only, whereas additionally adjusting for N-SES composite measures attenuated the effect estimates towards the null. This suggests that the neighborhood characteristics were potential confounders for the associations between air pollution and cognitive functioning and might lie on the biasing path that could not be blocked by controlling for only individual characteristics. The potential effect of neighborhood-level characteristics has been well documented on cognitive function, which might be more consequential to the elderly, as they spend longer time in neighborhoods due to the decreased mobility and rely more on community resources than younger populations (Glass & Balfour, 2003). A number of potential pathways by which lower N-SES neighborhoods impacts cognitive function have been proposed, including access to community resources (e.g., health service and facilities for physical activity) (Shih et al., 2011), social engagement and networking (Aneshensel, Ko, Chodosh, & Wight, 2011), cultural perception of illness and mental health (Wee et al., 2012), and exposure to environmental stressors.

Relatively fewer studies have investigated the potential effects of CO and NO_x_ on adult cognitive function, and the existing evidence remains controversial. Cullen et al. reported a significant association of residential exposure to ambient NO_x_ with better reaction time, a subdomain of cognition, with adjustment for individual characteristics, Townsend score, and population density, whereas the directions of associations with other subdomains, such as reasoning and numeric memory, were opposite but insignificant (Cullen et al., 2018). A study conducted in German women reported that NO_x_ was associated with lower cognitive performance as indicated by CERAD (Consortium to Establish a Registry for Alzheimer’s Disease) testing but not the Mini Mental State Examination (MMSE) (Schikowski et al., 2015), and another population-based study conducted in Germany found a significant association of exposure to NO_x_ with worse global cognition (Tzivian et al., 2017). Shin et al. observed a protective effect estimate of ambient CO on MMSE (Shin et al., 2019), and Li et al. did not observe a significant association between CO and the risk of vascular dementia among the Asian populations (C.-Y. Li, Li, Martini, & Hou, 2019). Although we observed a similar pattern for CO and NO_x_ in our study, the modeled components of air pollution were themselves highly correlated (PM_2.5_ vs. CO: Pearson’s *r* = 0.95; PM_2.5_ vs. NO_x_: *r* = 0.94). Therefore, we could not disentangle which of the pollutants were driving the observed association. Future studies are needed to disentangle the effects of highly correlated pollutants and understand their joint effects on cognition.

Our study showed that the associations of ambient CO, NO_x_, and PM_2.5_ with cognitive function were significantly modified by N-SES. The strongest associations between air pollution and cognitive function were found among participants assigned to three N-SES clusters (Clusters 4, 6, and 7). Participants living in disadvantaged neighborhoods (i.e., Cluster 6 and 7) were among the most vulnerable to the effects of air pollution on cognitive function, which is consistent with a previous study (J. Ailshire et al., 2017). Ailshire et al. found that the effect measure of PM_2.5_ associated with cognitive function was stronger among participants who were exposed to stressful neighborhood physical conditions (J. Ailshire et al., 2017). Also, they still observed a positive association between PM_2.5_ and cognitive function decline among participants with low neighborhood stress, which was consistent with our finding for Cluster 4 that had the highest N-SES among all clusters. Moreover, as Cluster 4 had the fewest vacant or rented households, the long-term exposure estimates might suffer less from exposure misclassification bias for the participants living in this cluster. We observed a significant protective effect of air pollution exposures on CFI for Cluster 2, which was opposite to our hypothesis and previous literature. To the best of our knowledge, only one previous study on air pollution and cognitive function has examined effect modification by N-SES. Wight et al. proposed that late-life cognition was a result of the interaction between individual characteristics and the environment given an interplay observed for individual and area-level education attainment (Wight et al., 2006). In addition, Gee and Payne-Sturges asserted that the community-level vulnerability resulting from low N-SES might translate to individual vulnerability to air pollution (Gee & Payne-Sturges, 2004). Our findings provide additional evidence for the interaction between N-SES and PM_2.5_ on cognitive function among older populations.

Our study has several limitations, which have to be considered when interpreting the results. First, the study was conducted based on an ongoing cohort study but only used the baseline data to perform a cross-sectional analysis because follow-up visits in this cohort are still on-going. This prevented us from establishing the temporal relationship between air pollution and cognitive function. Second, we assigned exposures as the 9-year average concentration for a fixed time period prior to the launch of the cohort in order to focus our analyses only on the variation in long-term spatial exposure; since participants were enrolled over time (between 2015-2019), the fixed exposure time period may contribute to some exposure measurement error by ignoring temporal variation. Third, since the long-term exposure was estimated at participants’ residences, and the information on residential addresses was collected at enrollment, information error for our exposure assessment might exist. To assess the potential impact of residential mobility, we performed a sensitivity analysis adjusting for neighborhood-level indicators of residence stability; this analysis demonstrated that our observed results were robust. Fourth, urban neighborhoods might experience gentrification that can change N-SES over time. While we matched the annual ACS data to participants based on their enrollment time, the snapshot of N-SES assigned to participants might not have fully represented the environment where participants lived. Finally, the study population might be not representative of the general Atlanta population in terms of gender (much higher proportion of females, as females are generally more likely to participate in scientific studies (Galea & Tracy, 2007)) and race (percent AA participants was much lower than the general population in Atlanta, GA, USA (U.S. Census Bureau, 2019)).

The most important strength of the current study is that, to our knowledge, it is the first study using two N-SES composite measures to adjust for confounding and effect modification by N-SES for the association between air pollution and cognitive function. Sixteen N-SES indicators were employed to create the N-SES composite measures, which guaranteed a good representation and identification of clusters of census tracts that shared a similar social, economic, and cultural context.

## CONCLUSIONS

This study emphasizes the necessity of considering N-SES as confounder and effect modifier when investigating the effect of ambient air pollution on cognitive function among the elderly. We demonstrated that N-SES and the long-term exposure to ambient air pollution could have a joint impact on cognitive function, and participants living in disadvantaged neighborhoods might be more susceptible to ambient air pollution than those living in other areas. This study identifies potentially vulnerable populations, those exposed to both relative higher concentrations of air pollution and disadvantaged social conditions, who may be at an increased risk from air pollution exposure on cognitive aging. Disproportionate exposure to air pollution and lower N-SES may play a critical role in leading to health disparities among marginalized populations. The investigation on the interaction of neighborhood features and environmental pollutants may offer important implications for the identification of health disparities in the context of environmental injustice.

## Data Availability

All data produced in the present study are available upon reasonable request to the authors.

## Notes

**Sources of support:** This work was based on information from the Emory Healthy Aging study, supported by HERCULES Pilot Project via National Institute of Environmental Health Sciences (NIEHS) P30ES019776 (PI: Anke Huels) and National Institute on Aging (NIA) R01AG070937 (PI: James J. Lah).

**Conflicts of interest:** The authors declare they have nothing to disclose.

### Competing Interest Statement

The authors have declared no competing interest.

### Funding Statement

This work was based on information from the Emory Healthy Aging study, supported by HERCULES Pilot Project via National Institute of Environmental Health Sciences (NIEHS) P30ES019776 (PI: Anke Huels) and National Institute on Aging (NIA) R01AG070937 (PI: James J. Lah).

### Author Declarations

IRB of Emory University gave ethical approval for this work.

## References

Abdi, H., & Williams, L. J. (2010). Principal component analysis. Wiley interdisciplinary reviews: computational statistics, 2(4), 433–459.

Ailshire, J., Karraker, A., & Clarke, P. (2017). Neighborhood social stressors, fine particulate matter air pollution, and cognitive function among older U.S. adults. Soc Sci Med, 172, 56–63. doi:10.1016/j.socscimed.2016.11.019

Ailshire, J. A., & Crimmins, E. M. (2014). Fine particulate matter air pollution and cognitive function among older US adults. American journal of epidemiology, 180(4), 359–366.

Amariglio, R. E., Donohue, M. C., Marshall, G. A., Rentz, D. M., Salmon, D. P., Ferris, S. H., … Sperling, R. A. (2015). Tracking early decline in cognitive function in older individuals at risk for Alzheimer disease dementia: the Alzheimer’s Disease Cooperative Study Cognitive Function Instrument. JAMA neurology, 72(4), 446–454.

Aneshensel, C. S., Ko, M. J., Chodosh, J., & Wight, R. G. (2011). The urban neighborhood and cognitive functioning in late middle age. J Health Soc Behav, 52(2), 163–179. doi:10.1177/0022146510393974

Basta, N. E., Matthews, F. E., Chatfield, M. D., Brayne, C., & Mrc, C. (2008). Community-level socio-economic status and cognitive and functional impairment in the older population. Eur J Public Health, 18(1), 48–54. doi:10.1093/eurpub/ckm076

Block, M. L., & Calderon-Garciduenas, L. (2009). Air pollution: mechanisms of neuroinflammation and CNS disease. Trends Neurosci, 32(9), 506–516. doi:10.1016/j.tins.2009.05.009

Bowe, B., Xie, Y., Yan, Y., & Al-Aly, Z. (2019). Burden of Cause-Specific Mortality Associated With PM2.5 Air Pollution in the United States. JAMA Netw Open, 2(11), e1915834. doi:10.1001/jamanetworkopen.2019.15834

Cullen, B., Newby, D., Lee, D., Lyall, D. M., Nevado-Holgado, A. J., Evans, J. J., … Cavanagh, J. (2018). Cross-sectional and longitudinal analyses of outdoor air pollution exposure and cognitive function in UK Biobank. Sci Rep, 8(1), 12089. doi:10.1038/s41598-018-30568-6

Delgado-Saborit, J. M., Guercio, V., Gowers, A. M., Shaddick, G., Fox, N. C., & Love, S. (2021). A critical review of the epidemiological evidence of effects of air pollution on dementia, cognitive function and cognitive decline in adult population. Sci Total Environ, 757, 143734. doi:10.1016/j.scitotenv.2020.143734

Espino, D. V., Lichtenstein, M. J., Palmer, R. F., & Hazuda, H. P. (2001). Ethnic differences in mini-mental state examination (MMSE) scores: where you live makes a difference. J Am Geriatr Soc, 49(5), 538–548. doi:10.1046/j.1532-5415.2001.49111.x

Fu, P., Guo, X., Cheung, F. M. H., & Yung, K. K. L. (2019). The association between PM2. 5 exposure and neurological disorders: a systematic review and meta-analysis. Science of the Total Environment, 655, 1240–1248.

Galea, S., & Tracy, M. (2007). Participation rates in epidemiologic studies. Ann Epidemiol, 17(9), 643–653. doi:10.1016/j.annepidem.2007.03.013

Gee, G. C., & Payne-Sturges, D. C. (2004). Environmental health disparities: a framework integrating psychosocial and environmental concepts. Environ Health Perspect, 112(17), 1645–1653. doi:10.1289/ehp.7074

Glass, T. A., & Balfour, J. L. (2003). Neighborhoods, aging, and functional limitations. Neighborhoods and health, 1, 303–334.

Goetz, M. E., Hanfelt, J. J., John, S. E., Bergquist, S. H., Loring, D. W., Quyyumi, A., … Lah, J. J. (2019). Rationale and Design of the Emory Healthy Aging and Emory Healthy Brain Studies. Neuroepidemiology, 53(3-4), 187–200. doi:10.1159/000501856

Guo, Y., Chan, C. H., Chang, Q., Liu, T., & Yip, P. S. F. (2019). Neighborhood environment and cognitive function in older adults: A multilevel analysis in Hong Kong. Health Place, 58, 102146. doi:10.1016/j.healthplace.2019.102146

Hicken, M. T., Adar, S. D., Hajat, A., Kershaw, K. N., Do, D. P., Barr, R. G., … Diez Roux, A. V. (2016). Air Pollution, Cardiovascular Outcomes, and Social Disadvantage: The Multiethnic Study of Atherosclerosis. Epidemiology, 27(1), 42–50. doi:10.1097/EDE.0000000000000367

Kodinariya, T. M., & Makwana, P. R. (2013). Review on determining number of Cluster in K-Means Clustering. International Journal, 1(6), 90–95.

Krieger, N., Chen, J. T., Waterman, P. D., Soobader, M.-J., Subramanian, S., & Carson, R. J. A. j. o. e. (2002). Geocoding and monitoring of US socioeconomic inequalities in mortality and cancer incidence: does the choice of area-based measure and geographic level matter? the Public Health Disparities Geocoding Project. 156(5), 471–482.

Lang, I. A., Llewellyn, D. J., Langa, K. M., Wallace, R. B., Huppert, F. A., & Melzer, D. (2008). Neighborhood deprivation, individual socioeconomic status, and cognitive function in older people: analyses from the English Longitudinal Study of Ageing. J Am Geriatr Soc, 56(2), 191–198. doi:10.1111/j.1532-5415.2007.01557.x

Li, C.-Y., Li, C.-H., Martini, S., & Hou, W.-H. (2019). Association between air pollution and risk of vascular dementia: A multipollutant analysis in Taiwan. Environment international, 133, 105233.

Li, C., Neugroschl, J., Luo, X., Zhu, C., Aisen, P., Ferris, S., & Sano, M. (2017). The Utility of the Cognitive Function Instrument (CFI) to Detect Cognitive Decline in Non-Demented Older Adults. J Alzheimers Dis, 60(2), 427–437. doi:10.3233/JAD-161294

Lin, H., Guo, Y., Zheng, Y., Zhao, X., Cao, Z., Rigdon, S. E., … Wu, F. (2017). Exposure to ambient PM2.5 associated with overall and domain-specific disability among adults in six low-and middle-income countries. Environ Int, 104, 69–75. doi:10.1016/j.envint.2017.04.004

Maantay, J. (2002). Mapping environmental injustices: pitfalls and potential of geographic information systems in assessing environmental health and equity. Environ Health Perspect, 110 Suppl 2, 161–171. doi:10.1289/ehp.02110s2161

Messer, L. C., Laraia, B. A., Kaufman, J. S., Eyster, J., Holzman, C., Culhane, J., … O’Campo, P. (2006). The development of a standardized neighborhood deprivation index. J Urban Health, 83(6), 1041–1062. doi:10.1007/s11524-006-9094-x

Ou, C. Q., Hedley, A. J., Chung, R. Y., Thach, T. Q., Chau, Y. K., Chan, K. P., … Lam, T. H. (2008). Socioeconomic disparities in air pollution-associated mortality. Environ Res, 107(2), 237–244. doi:10.1016/j.envres.2008.02.002

Schikowski, T., Vossoughi, M., Vierkotter, A., Schulte, T., Teichert, T., Sugiri, D., … Luckhaus, C. (2015). Association of air pollution with cognitive functions and its modification by APOE gene variants in elderly women. Environ Res, 142, 10–16. doi:10.1016/j.envres.2015.06.009

Senthilkumar, N., Gilfether, M., Metcalf, F., Russell, A. G., Mulholland, J. A., & Chang, H. H. (2019). Application of a Fusion Method for Gas and Particle Air Pollutants between Observational Data and Chemical Transport Model Simulations Over the Contiguous United States for 2005-2014. Int J Environ Res Public Health, 16(18). doi:10.3390/ijerph16183314

Sheffield, K. M., & Peek, M. K. (2009). Neighborhood context and cognitive decline in older Mexican Americans: results from the Hispanic Established Populations for Epidemiologic Studies of the Elderly. Am J Epidemiol, 169(9), 1092–1101. doi:10.1093/aje/kwp005

Shih, R. A., Ghosh-Dastidar, B., Margolis, K. L., Slaughter, M. E., Jewell, A., Bird, C. E., … Espeland, M. A. (2011). Neighborhood socioeconomic status and cognitive function in women. Am J Public Health, 101(9), 1721–1728. doi:10.2105/AJPH.2011.300169

Shin, J., Han, S. H., & Choi, J. (2019). Exposure to Ambient Air Pollution and Cognitive Impairment in Community-Dwelling Older Adults: The Korean Frailty and Aging Cohort Study. Int J Environ Res Public Health, 16(19). doi:10.3390/ijerph16193767

Tallon, L. A., Manjourides, J., Pun, V. C., Salhi, C., & Suh, H. (2017). Cognitive impacts of ambient air pollution in the National Social Health and Aging Project (NSHAP) cohort. Environ Int, 104, 102–109. doi:10.1016/j.envint.2017.03.019

Tzivian, L., Jokisch, M., Winkler, A., Weimar, C., Hennig, F., Sugiri, D., … Heinz Nixdorf Recall Study, G. (2017). Associations of long-term exposure to air pollution and road traffic noise with cognitive function-An analysis of effect measure modification. Environ Int, 103, 30–38. doi:10.1016/j.envint.2017.03.018

U.S. Census Bureau. (2019). 2019: ACS 1-Year Estimates Detailed Tables. Retrieved from https://data.census.gov/cedsci/table?q=atlanta%20race&tid=ACSDT1Y2019.B02001&hidePreview=false

US Department of Health and Human Services. (2020). 2021 Poverty guidelines for the 48 contiguous States and the District of Columbia. Retrieved from https://dch.georgia.gov/federal-poverty-guidelines-0

Walker, K., Eberwein, K., & Herman, M. J. (2018). Tidycensus: Load us census boundary and attribute data as’ tidyverse’and’sf’-ready data frames. R package version 0.10.2.

Wee, L. E., Yeo, W. X., Yang, G. R., Hannan, N., Lim, K., Chua, C., … Shen, H. M. (2012). Individual and Area Level Socioeconomic Status and Its Association with Cognitive Function and Cognitive Impairment (Low MMSE) among Community-Dwelling Elderly in Singapore. Dement Geriatr Cogn Dis Extra, 2(1), 529–542. doi:10.1159/000345036

Wight, R. G., Aneshensel, C. S., Miller-Martinez, D., Botticello, A. L., Cummings, J. R., Karlamangla, A. S., & Seeman, T. E. (2006). Urban neighborhood context, educational attainment, and cognitive function among older adults. Am J Epidemiol, 163(12), 1071–1078. doi:10.1093/aje/kwj176

Wingo, A. P., Wingo, T. S., Fan, W., Bergquist, S., Alonso, A., Marcus, M., … Lah, J. J. (2020). Purpose in life is a robust protective factor of reported cognitive decline among late middle-aged adults: The Emory Healthy Aging Study. J Affect Disord, 263, 310–317. doi:10.1016/j.jad.2019.11.124

Yu, H. F., Russell, A., Mulholland, J., Odman, T., Hu, Y. T., Chang, H. H., & Kumar, N. (2018). Cross-comparison and evaluation of air pollution field estimation methods. Atmospheric Environment, 179, 49–60. doi:10.1016/j.atmosenv.2018.01.045

