## Supplemental Materials for "Environmental injustice - Neighborhood characteristics as confounders and effect modifiers for the association between air pollution exposure and cognitive function"

**Sources of support:** This work was based on information from the Emory Healthy Aging study, supported by HERCULES Pilot Project via National Institute of Environmental Health Sciences (NIEHS) P30ES019776 (PI: Anke Huels) and National Institute on Aging (NIA) R01AG070937 (PI: James J. Lah).

### Supplementary Materials

**Table S1.** Description of the 9-year averaged exposure to air pollutants estimated by CMAQ-RLINE model.

| Characteristics | CO (ppb) | | | $NO_x$ (ppb) | | | $PM_{2.5}$ ($\mu g/m^3$) | | |
|---|---|---|---|---|---|---|---|---|---|
| | Median | IQR | Range | Median | IQR | Range | Median | IQR | Range |
| Age | | | | | | | | | |
| Mean (SD) | | 0.021 [a] | | | 0.017 [a] | | | 0.031 [a] | |
| Gender | | | | | | | | | |
| Female | 661.5 | 321.1 | 4,488.5 | 46.6 | 27.5 | 415.1 | 14.5 | 1.3 | 13.4 |
| Male | 686.8 | 345.7 | 4,474.9 | 48.1 | 29.0 | 413.2 | 14.7 | 1.3 | 15.1 |
| Race | | | | | | | | | |
| White/Caucasian | 677.7 | 336.0 | 4,488.5 | 47.0 | 28.5 | 415.1 | 14.6 | 1.3 | 13.4 |
| Black/African American | 625.9 | 276.9 | 4,453.6 | 48.2 | 25.7 | 410.0 | 14.3 | 1.0 | 15.0 |
| Others [b] | 650.9 | 286.3 | 2,069.4 | 45.0 | 24.7 | 189.8 | 14.5 | 1.1 | 7.2 |
| Hispanic | | | | | | | | | |
| No | 669.1 | 327.6 | 4,488.5 | 47.0 | 27.9 | 415.1 | 14.6 | 1.3 | 15.3 |
| Yes | 655.6 | 345.6 | 2,116.6 | 46.1 | 29.0 | 191.6 | 14.5 | 1.4 | 7.3 |
| Household income | | | | | | | | | |
| USD < 50,000 | 681.1 | 319.9 | 3,424.7 | 49.8 | 28.2 | 286.0 | 14.6 | 1.3 | 9.3 |
| USD 50,000-100,000 | 651.9 | 320.2 | 4,480.0 | 45.2 | 26.6 | 412.9 | 14.4 | 1.3 | 15.1 |
| USD >= 100,000 | 677.6 | 337.3 | 4,488.5 | 47.1 | 28.6 | 412.9 | 14.6 | 1.3 | 13.4 |
| Highest education | | | | | | | | | |
| High school or less | 605.9 | 311.6 | 2,849.5 | 42.7 | 27.6 | 264.0 | 14.3 | 1.2 | 8.2 |
| Some college credit, but no degree | 602.9 | 298.3 | 4,469.1 | 42.2 | 25.2 | 412.9 | 14.3 | 1.3 | 13.2 |
| Associate degree | 596.4 | 285.6 | 3,061.0 | 41.8 | 25.1 | 284.1 | 14.2 | 1.3 | 9.3 |
| Bachelor degree | 670.3 | 321.1 | 4,488.5 | 46.8 | 26.7 | 412.5 | 14.6 | 1.2 | 13.4 |
| Master degree | 697.0 | 320.7 | 4,453.6 | 49.8 | 27.9 | 415.1 | 14.8 | 1.3 | 15.1 |
| Professional or doctorate degree | 751.9 | 320.7 | 4,474.9 | 54.6 | 28.6 | 411.3 | 15.0 | 1.3 | 13.1 |
| Mild cognitive impairment | | | | | | | | | |
| No | 669.9 | 326.5 | 4,488.5 | 47.0 | 27.9 | 415.1 | 14.6 | 1.3 | 15.3 |
| Yes | 658.9 | 328.3 | 1,882.9 | 47.4 | 30.2 | 163.8 | 14.5 | 1.4 | 6.9 |
| Alzheimer's disease | | | | | | | | | |
| No | 668.7 | 326.9 | 4,488.5 | 47.0 | 27.9 | 415.1 | 14.5 | 1.3 | 15.3 |
| Yes | 668.0 | 338.7 | 1,673.7 | 47.4 | 31.9 | 151.8 | 14.6 | 1.4 | 6.4 |
| Other dementia | | | | | | | | | |
| No | 668.7 | 326.4 | 4,488.5 | 47.0 | 27.8 | 415.1 | 14.5 | 1.3 | 15.3 |
| Yes | 657.3 | 348.8 | 1,631.3 | 47.4 | 33.5 | 151.8 | 14.4 | 1.4 | 6.5 |

[a] Pearson's correlation coefficient.
[b] Others included Asian, American Indian, Alaska Native, Native Hawaiian and other pacific islander, and multi-race.

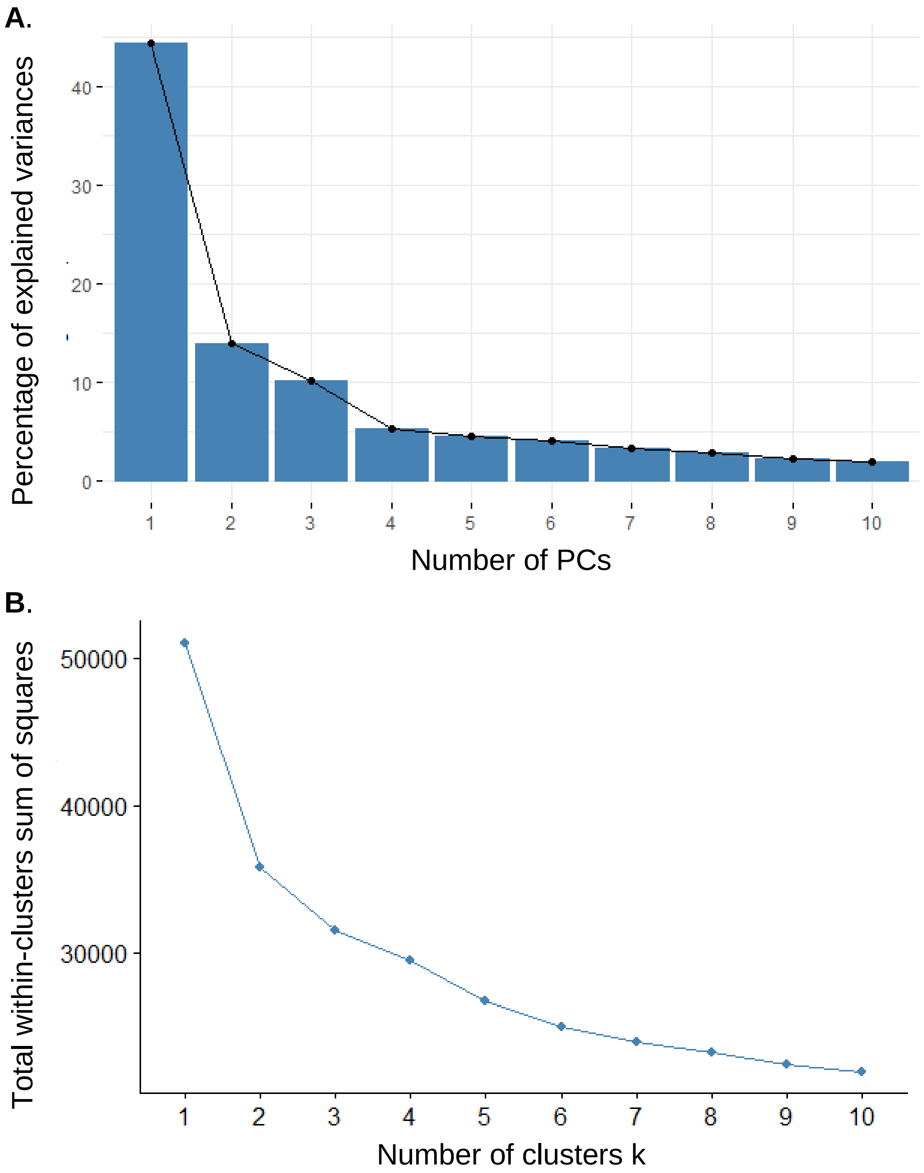


**Figure S1**. The diagnostic plots of principal component analysis and k-means clustering analysis on 16 neighborhood-level socioeconomic status indicators. (A) Scree plot of principal component (PC) analysis on 16 N-SES indicators. (B) Determination of optimal clusters for k-means clustering analysis by Elbow method.

**Table S2.** Associations between 9-year averaged exposure to ambient air pollution and Cognitive Function Instrument stratified by the N-SES clusters.

| Pollutants | Cluster index | Effect estimates[a] | 95% Confidence interval | |
|---|---|---|---|---|
| | | | Lower | Upper |
| CO | 1 | 3.3 | -2.1 | 8.9 |
| | 2 | -4.7 | -8.5 | -0.8 |
| | 3 | -1.2 | -3.9 | 1.5 |
| | 4 | 6.3 | 2.1 | 10.8 |
| | 5 | -4.1 | -9.1 | 1.3 |
| | 6 | 5.4 | -0.2 | 11.4 |
| | 7 | 13.4 | 1.3 | 26.9 |
| $NO_x$ | 1 | 2.9 | -2.4 | 8.4 |
| | 2 | -4.3 | -7.8 | -0.6 |
| | 3 | -1 | -3.6 | 1.7 |
| | 4 | 7.2 | 2.9 | 11.7 |
| | 5 | -4.1 | -9.5 | 1.6 |
| | 6 | 4.9 | -0.4 | 10.4 |
| | 7 | 13.4 | 0.3 | 28.2 |
| $PM_{2.5}$ | 1 | 2.2 | -4.3 | 9.2 |
| | 2 | -6.5 | -11.5 | -1.1 |
| | 3 | -1.4 | -5.2 | 2.6 |
| | 4 | 7.3 | 2.2 | 12.6 |
| | 5 | -4.9 | -10.1 | 0.6 |
| | 6 | 9.8 | 2.2 | 18 |
| | 7 | 17.6 | 2.8 | 34.5 |

[a] Percent change (%) of CFI associated with an interquartile range increase (CO, 328.4ppb; $NO_x$, 28.0ppb; $PM_{2.5}$, 1.3μg/$m^3$) in the exposure estimate.

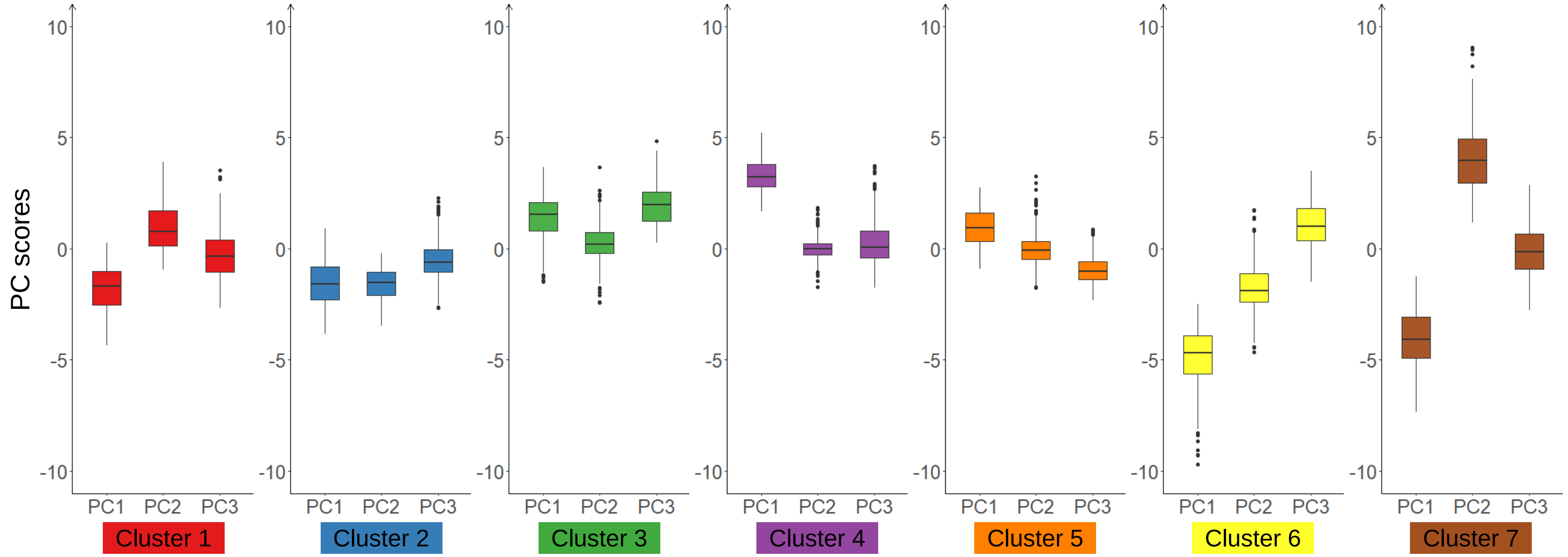


**Figure S2**. The distribution of the first, second, and third principal components (PC1, PC2, and PC3) across the seven cluster

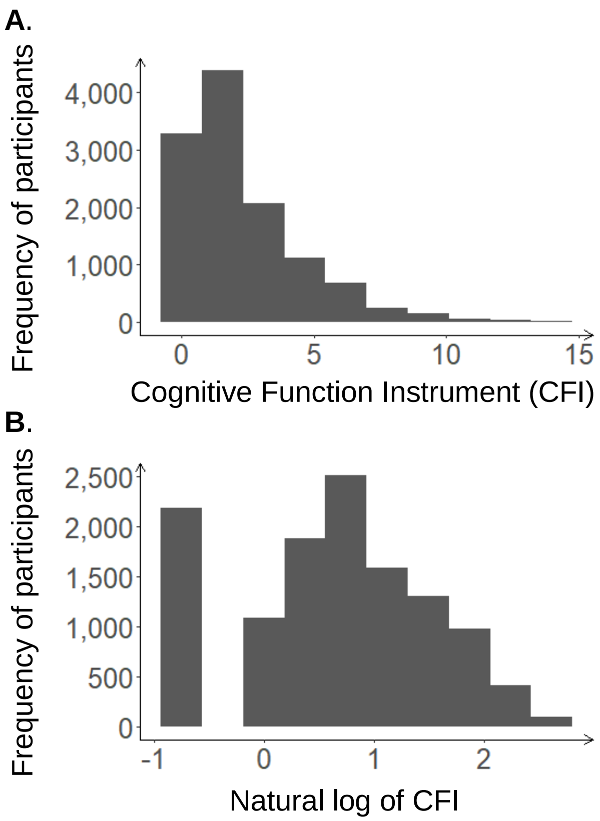


**Figure S3.** The distribution of Cognitive Function Instrument and its natural log for all the participants

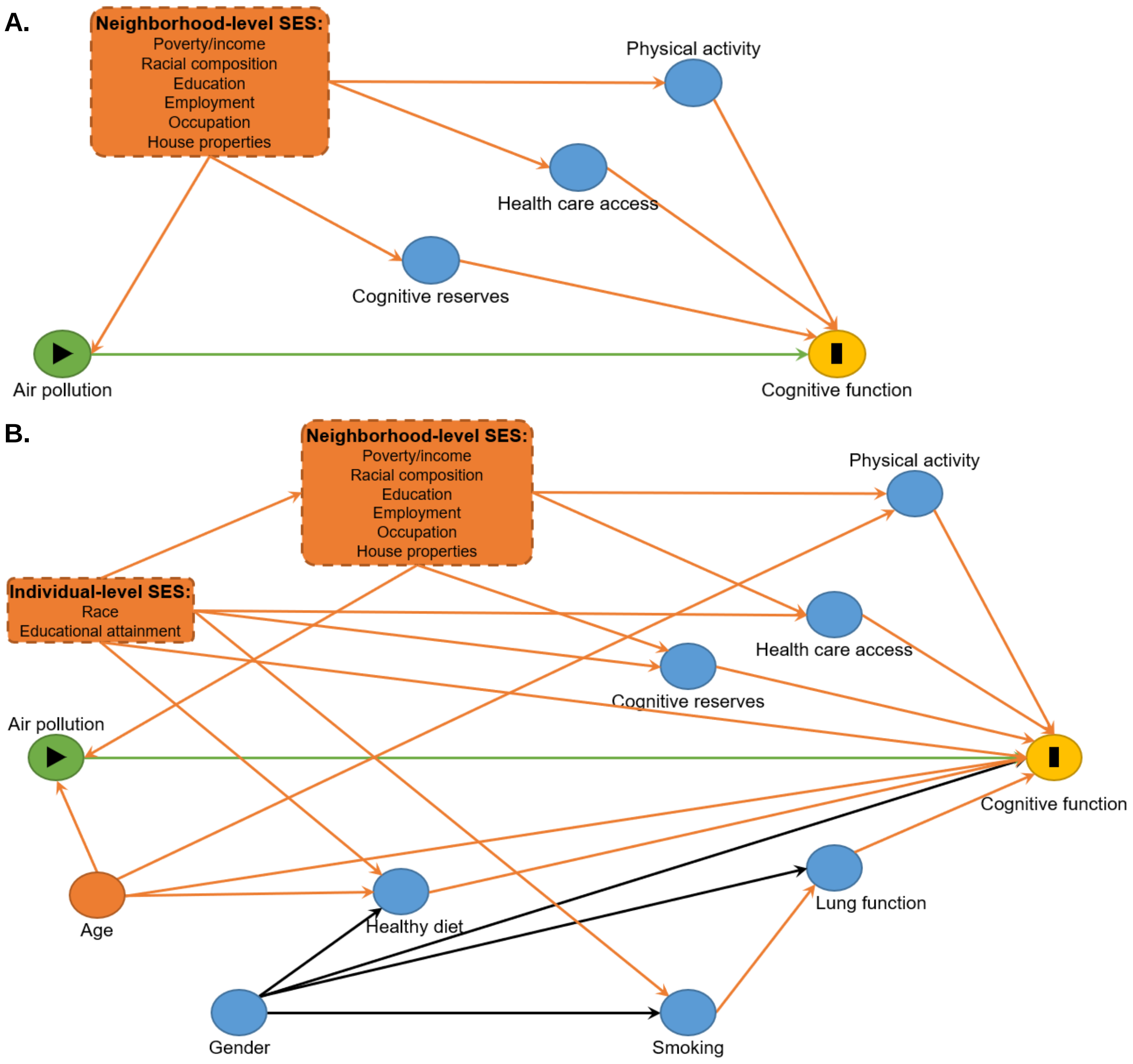


**Figure S4.** Directed acyclic graphs of the hypothesized causal structure linking air pollution and cognitive function in the current analysis. (A) Assessment of the confounding of neighborhood-level socioeconomic status. (N-SES) on the association between air pollution and cognitive function. (B) Identification of the minimal sufficient adjustment set for estimating the total effect of air pollution on cognitive function. Circle with black triangle in the center represents the exposure; circle with black I in the center represents the outcome. All other circles and blocks represent confounders and mediators. Unidirectional-pointed arrows represents causal effects. The biasing paths and the minimal sufficient set of confounders for identifying the total effect of ambient air pollution on cognitive function are colored in orange. Individual-level SES was indicated by race and educational attainment. Sixteen N-SES indicators were collected belonging to the six domains: poverty/income, racial composition, education, employment, occupation, housing properties, and the complex intercorrelation among the indicators was ignored for simplification. Correspondingly, a N-SES composite measure was generated to serve as the confounder in models.

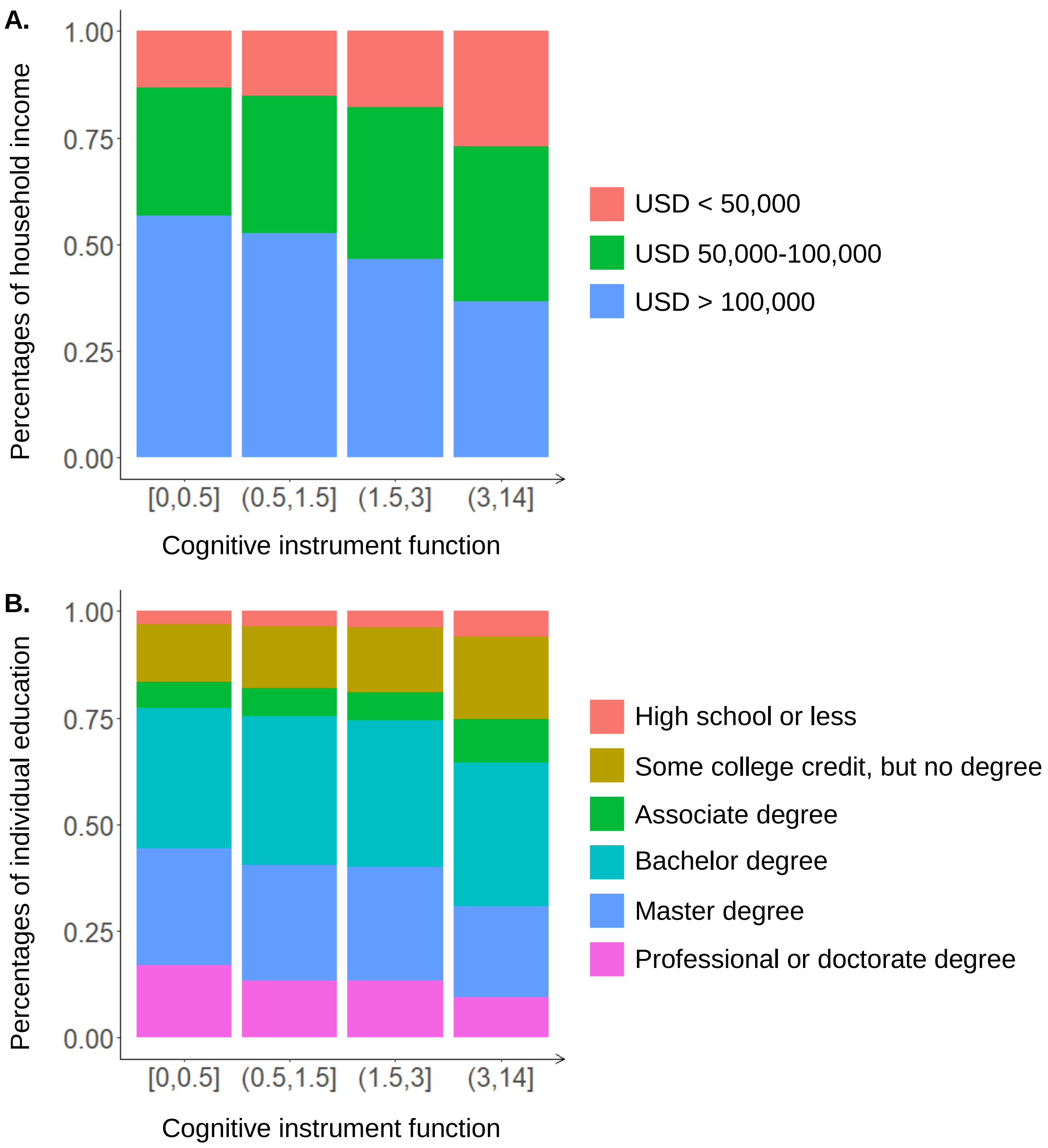


**Figure S5.** The distribution of individual education and household income for the study population by cognitive instrument function (CFI). The CFI were stratified by quartiles.

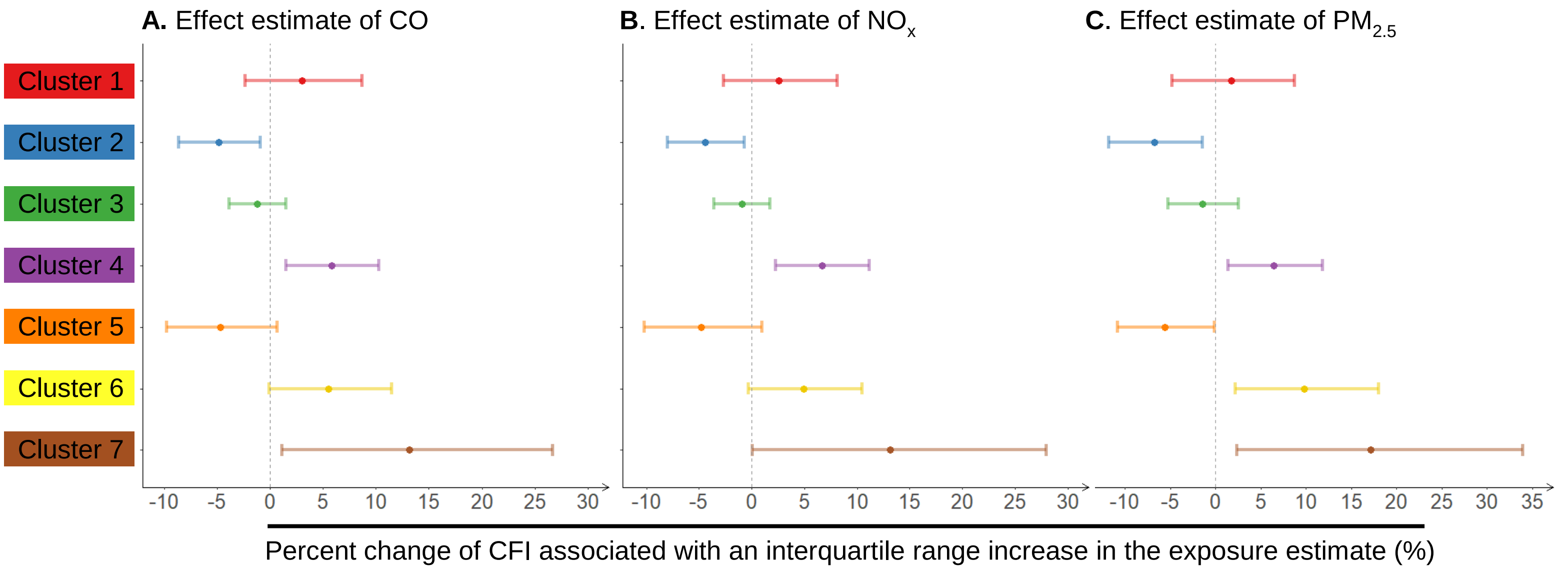


**Figure S6.** The associations between 9-year averaged exposure to ambient air pollution and Cognitive Function Instrument (CFI) the modification of neighborhood socioeconomic status (N-SES) among participants over 50, living in Metro Atlanta, and enrolled in Emory Healthy Aging Study, 2014-2020. The results were generated based on Model 6. Compared to Model 5, the inclusion of residential stability in Model 6 did not impact observed association between air pollution and CFI with the modification of N-SES. X-axis represented the percent change of CFI associated with an interquartile range increase in the exposure estimate. The cluster-specific effect estimates were colored in the same way as the N-SES map in Figure 1A.
